# Trends and variation in prescribing of suboptimal statin treatment regimes: a cohort study in English primary care

**DOI:** 10.1101/19008748

**Authors:** Helen J Curtis, Alex J Walker, Brian MacKenna, Richard Croker, Ben Goldacre

## Abstract

**Objectives:** We set out to describe trends and variation in statin prescribing in England that breaches 2014 national guidance on “high-intensity” statins. We identify factors associated with breaching; and assess the feasibility of rapid prescribing behaviour change.

**Design, Setting and Participants:** Retrospective cohort study in NHS primary care in England, including all 8,142 standard general practices from August 2010 to March 2019.

**Main Outcome Measures:** We categorised statins as high or low/medium-intensity based on two different thresholds, and calculated the proportion prescribed below these thresholds across all practices. We plotted trends and geographical variation, carried out mixed effects logistic regression to identify practice characteristics associated with breaching guidance, and used indicator saturation to identify practices exhibiting sudden changes in prescribing.

**Results:** We included all 8,142 practices across the study period. The proportion of statin prescriptions below the recommended 40% LDL-lowering threshold decreased gradually since 2012 from 80% to 45%; the proportion below a pragmatic 37% threshold decreased from 30% to 18%. The 2014 guidance had minimal impact on these trends. We found wide variation between practices (interdecile ranges 20% to 85% and 10% to 30% respectively in 2018). Mixed effects logistic regression did not identify practice characteristics strongly associated with breaching guidance. Indicator saturation identified several practices exhibiting sudden changes in prescribing towards greater guideline compliance.

**Conclusions:** Breaches of English guidance on choice of statin remain common, with substantial variation between GP practices. Some practices and regions have implemented rapid change, indicating the feasibility of rapid prescribing behaviour change. We discuss the potential for a national strategic approach, using data and evidence to optimise care, including targeted education alongside audit and feedback to outliers through services such as OpenPrescribing.

**Summary:** *What is already known on this topic:* English national guidance recommends the use of a high-intensity statin, capable of reducing LDL (low-density lipoprotein) cholesterol by 40% or more. Adherence at the time of guideline release was low, but has not been documented since.

*What this study adds:* Adherence is improving, but breaches of national guidance remain common, with 45% of prescriptions below the recommended strength, and there is very substantial variation between practices. Some practices have exhibited rapid positive change in prescribing, which indicates that better adherence could readily be achieved. We have produced a live data tool allowing anyone to explore any practice’s current statin prescribing behaviour.

## Introduction

Statins are very widely used to control serum cholesterol and reduce the risk of cardiovascular disease (CVD), with up to 7 million of the UK population (64.6 million) taking them in 2014 [1]. This makes statins the most commonly prescribed class of drugs in England, with 72.5 million prescriptions costing over £200m dispensed during 2017 [2,3]. The 2014 guidance on lipid modification by the National Institute for Health and Care Excellence (NICE) [4] recommends the use of a high-intensity statin, capable of reducing LDL (low-density lipoprotein) cholesterol by 40% or more [5]. This recommendation was made on the basis that higher intensity treatment offers substantially greater reduction in cardiovascular risk, at similar risk and cost. Similar guidelines are in place across many other countries. The high-intensity treatment options available in the UK are: atorvastatin 20mg and above; simvastatin 80mg; and rosuvastatin 10mg and above. Fluvastatin is medium-intensity at its highest dose; and pravastatin is low-intensity at all doses.

Two retrospective analysis studies carried out in UK patient-level datasets indicate that a huge shift in treatment would be required to meet the new recommendations: in 2013, only 24% of patients with CVD were receiving high-intensity statins [6]; and in 2014, 31% of patients with atherosclerotic CVD received high-intensity statin, with 21% not receiving statins at all [7]. This second study also noted that only 6% of these CVD patients were receiving statin therapy fully in line with the new guidelines for secondary prevention (atorvastatin 80 mg or equivalent); similarly, for patients in the high risk category for CVD, only 15% were on high-intensity statins (minimum atorvastatin 20mg or equivalent).

Our group runs OpenPrescribing.net, an online service that gives free and open access to monthly prescriptions data and charts describing various treatment choices at every general practice in England, with over 130,000 unique users during the past year. This service includes a standard “audit and feedback” measure that describes the prescribing of low/medium-intensity statins, as a proportion of all statin prescribing, at each practice ([link](https://openprescribing.net/measure/statinintensity/)). We were concerned to note, anecdotally, that breaches of NICE guidance on statin prescribing were extremely prevalent. We therefore set out to describe trends and variation in the proportion of all statin prescribing in English primary care that breaches this guidance; to identify factors associated with breaching; and to assess the feasibility of prescribing behaviour change by ascertaining whether there were individual practices that had rapidly implemented substantial changes.

## Methods

### Study design

A retrospective cohort study of statin prescribing behaviour using routinely collected primary care prescribing data. Outcomes were not pre-specified.

### Setting

NHS primary care in England, including all standard general practices with statin prescriptions dispensed from August 2010 to March 2019.

### Data sources

Monthly practice-level data on all items prescribed in NHS primary care in England and dispensed in the community is published by the NHS Business Services Authority (BSA). We used data from OpenPrescribing.net, which imports this data, alongside various other datasets giving practice characteristics, as previously described [8,9]. National prescribing data record the number of items of each individual drug presentation prescribed by every practice in England, for every month since August 2010 [10]. Prescribing activity for each drug presentation is measured as the number of *items* (corresponding to the number of prescriptions dispensed) and *quantity* (the total number of tablets/millilitres etc). Treatment duration and dosing regimen cannot strictly be ascertained, but statin treatment is typically a single tablet once a day, meaning that tablet strengths are an appropriate surrogate for statin dose.

### Data processing

We extracted all available monthly data from August 2010 to March 2019 inclusive for all statins (Appendix 1). We restricted our analysis to standard general practices (setting code “4”), excluding atypical settings such as prisons and out-of-hours services, using NHS Digital organisation data [11]. For analysis of current prescribing, we restricted data to 2018. We obtained the number of patients registered per practice from NHS Digital [12]. We classified statins according to their strength, with all tablets of rosuvastatin <10mg, atorvastatin <20mg and simvastatin <80mg classed as low and medium-intensity according to NICE guidelines, and we also implemented a more pragmatic classification based on a statin intensity threshold of 37%, where Rosuvastatin 5mg, Atorvastatin 10mg and Simvastatin 40mg were grouped with the high-intensity formulations (BNF codes in Appendix 1). When using quantity we excluded liquid and other non-tablet formulations.

### National trends

Across all practices we summed the total statin items prescribed per month, the proportion of which were of low and medium-intensity, and the rate of low and medium-intensity statins prescribed per 1,000 registered patients, and plotted these as time trends charts.

### CCG-level variation

Every practice belongs to a regional clinical commissioning group (CCG), which typically contain 20-100 practices. We calculated the proportion of low and medium-intensity statin prescribing for each CCG in 2018, and displayed the results as choropleth maps.

### Practice-level variation

We calculated the prescribing rate per 1,000 registered patients in each practice for all statins, and for each of the main subgroups. We displayed the data as deciles and centiles on time trends charts. We repeated this for the proportion of statin items prescribed in low and medium-intensity formulations, and for the proportion of all statin tablets (quantity) prescribed as specific atorvastatin and simvastatin formulations (all presentations which were not tablets or capsules were excluded).

### Logistic regression

We created a mixed effects logistic regression model to assess the factors associated with a practice prescribing higher proportions of low and medium-intensity statins in 2018. The fixed effect variables, selected *a priori* on the basis of clinical interest and data availability, were as follows: proportion of patients registered aged over 65; proportion of patients with a long-term health condition; index of multiple deprivation; Quality Outcomes Framework (QOF) score [13]; practice list size (NHS Digital); and rural/urbanness of practice postcode [14]. Practices with missing data were dropped from that part of the analysis. Continuous variables were categorised into quintiles to allow for nonlinearity of effects and to improve the intelligibility of results. We also included CCG as a random effect to assess the extent to which CCG membership explained variation in prescribing behaviour. The main outcome used was low- and medium-intensity statin prescriptions as a proportion of all statin prescriptions. This proportion was transformed using a conditional logit transformation [15]. This can be conceived of as a logistic regression analysis where each prescription written by each GP is a binary choice to give either compliant or non-compliant treatment. The model was used to calculate odds ratios and 95% confidence intervals (CI) for each of the fixed effect variables, as well as an R-squared value (along with the significance level) to describe the degree of variance associated with CCG membership.

### Practices that have changed quickly

In order to identify practices that had changed their statin dose prescribing behaviour rapidly, we calculated the proportion of low- and medium-intensity statin prescribing, for each month, for each practice. Using these practice-level time series, we applied our previously described indicator saturation method [16], to detect the timing, slope and magnitude of changes in prescribing. From these results, we then filtered to find practices that had a large total change and those with a rapid slope of change. Examples of rapidly changing practices were plotted on a line graph.

### Software and Reproducibility

Data management was performed using Python and Google BigQuery, with analysis carried out using Stata 14.2 / Python. All data is shared openly online alongside all code for data management and analysis: https://github.com/ebmdatalab/statins-dose-paper.

### Ethical approval

This study uses exclusively open, publicly available data, therefore no ethical approval was required.

## Results

### Study population

All 8,142 standard general practices in England were included across the whole time period. In 2018 there were 7,210, organised into 195 local CCGs.

### National trends

The overall rate of prescribing of statins in England has increased, from around 85-90 items per thousand patients per month in 2011/12 to around 100 items in 2018/19, but with the rate of increase slowing over time (Figure 1a). Low and medium-intensity statins, using the NICE criteria of 40% reduction in LDL cholesterol, consistently made up 80% of statin prescriptions in 2011-12; this decreased steadily from late 2012 at a rate of 5.4 percentage points per year, approaching 45% of all statin prescriptions in 2019 (Figure 1b). The proportion of statins prescribed in formulations giving <37% reduction is also consistently declining, from a peak of 30% in 2013 to 18% in 2019. eFigure 1 shows the time trends in the most common formulations of statins as a proportion of all statin prescribing across England. Notably, prescribing of atorvastatin 10mg, 20mg and 40mg and simvastatin 20mg underwent a sharp increase in 2012, coinciding with a rapid reduction in simvastatin 40mg. Thereafter, prescribing of high-intensity atorvastatin (20mg-80mg) increased, while atorvastatin 10mg levelled off and all simvastatin declined, including the high-intensity form (80mg).

**Figure 1.**
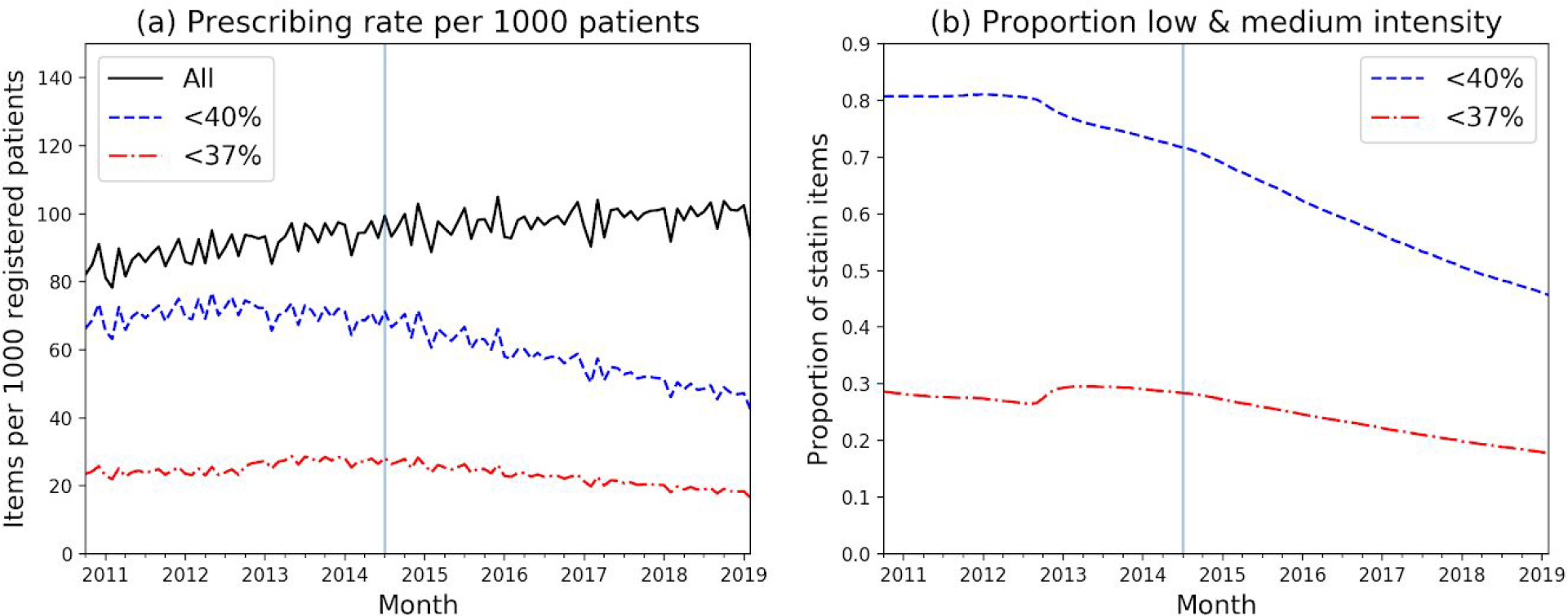
Monthly statin prescribing across English NHS practices. (a) Total statin items prescribed per 1000 registered patients, and those of low and medium-intensity, at both 40% and 37% intensity thresholds; (b) the proportion of statin items prescribed which were of low and medium-intensity, including both intensity thresholds. Vertical line indicates release of NICE guidance, July 2014.

### CCG-level variation in statin prescribing

Among England’s local practice groups, CCGs, the proportion of statins prescribed in low intensity formulations in 2018 ranged widely, from approximately 25-65%, or 7-31% under the 37% threshold (Figure 2). There was little notable geographic clustering, except that the lowest figures (closest to NICE recommendations) are in central London and one northern region (Bradford area).

**Figure 2.**
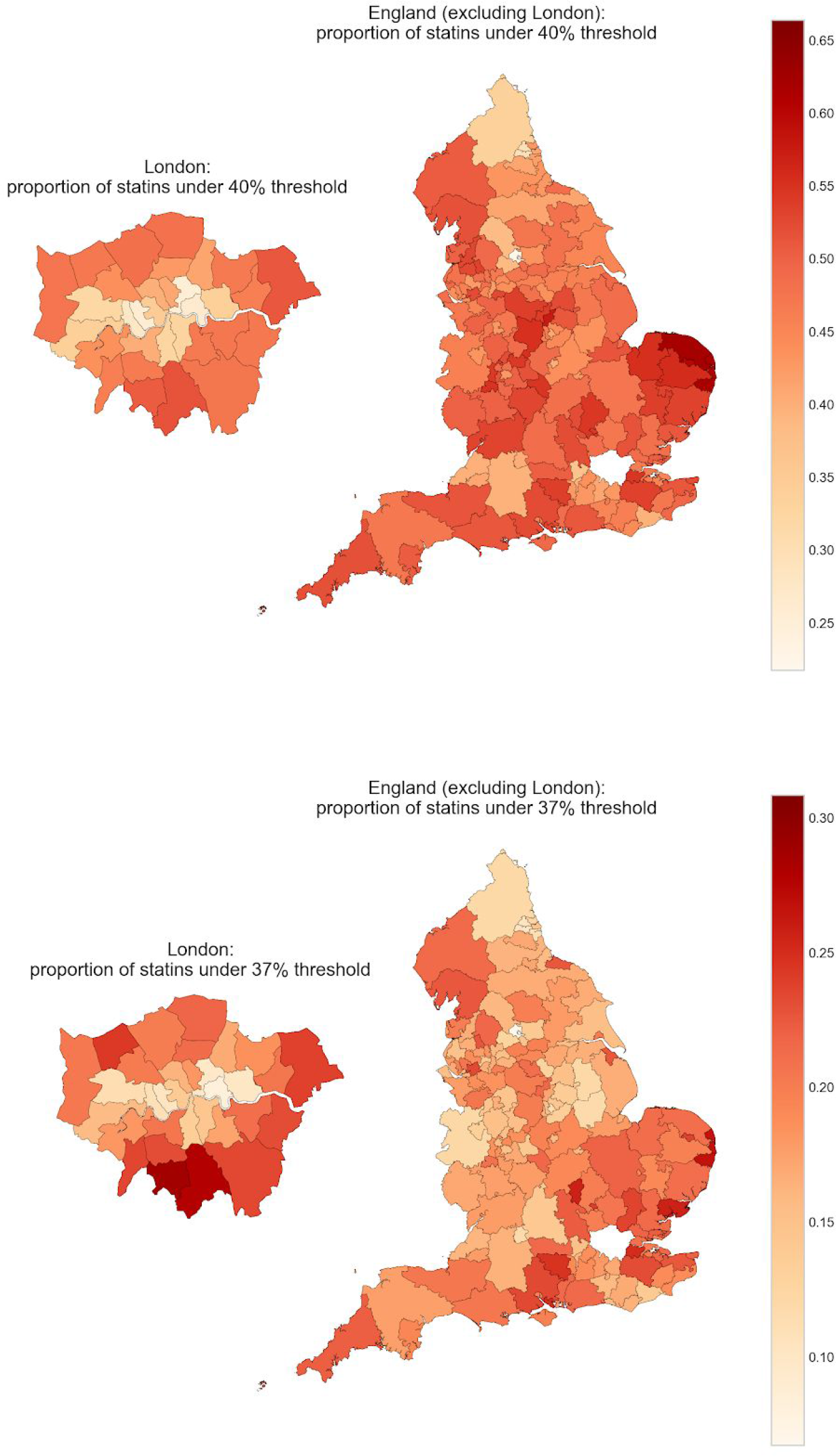
The proportion of all statin items prescribed in low and medium-intensity formulations (<40%/<37%) across each CCG in England, 2018.

### Practice-level variation in statin prescribing

The national decline in the proportion of low and medium-intensity statin items since 2012 was reflected across all deciles at the level of individual GP practices, but variation between practices increased slightly over time (Figure 3a).

**Figure 3.**
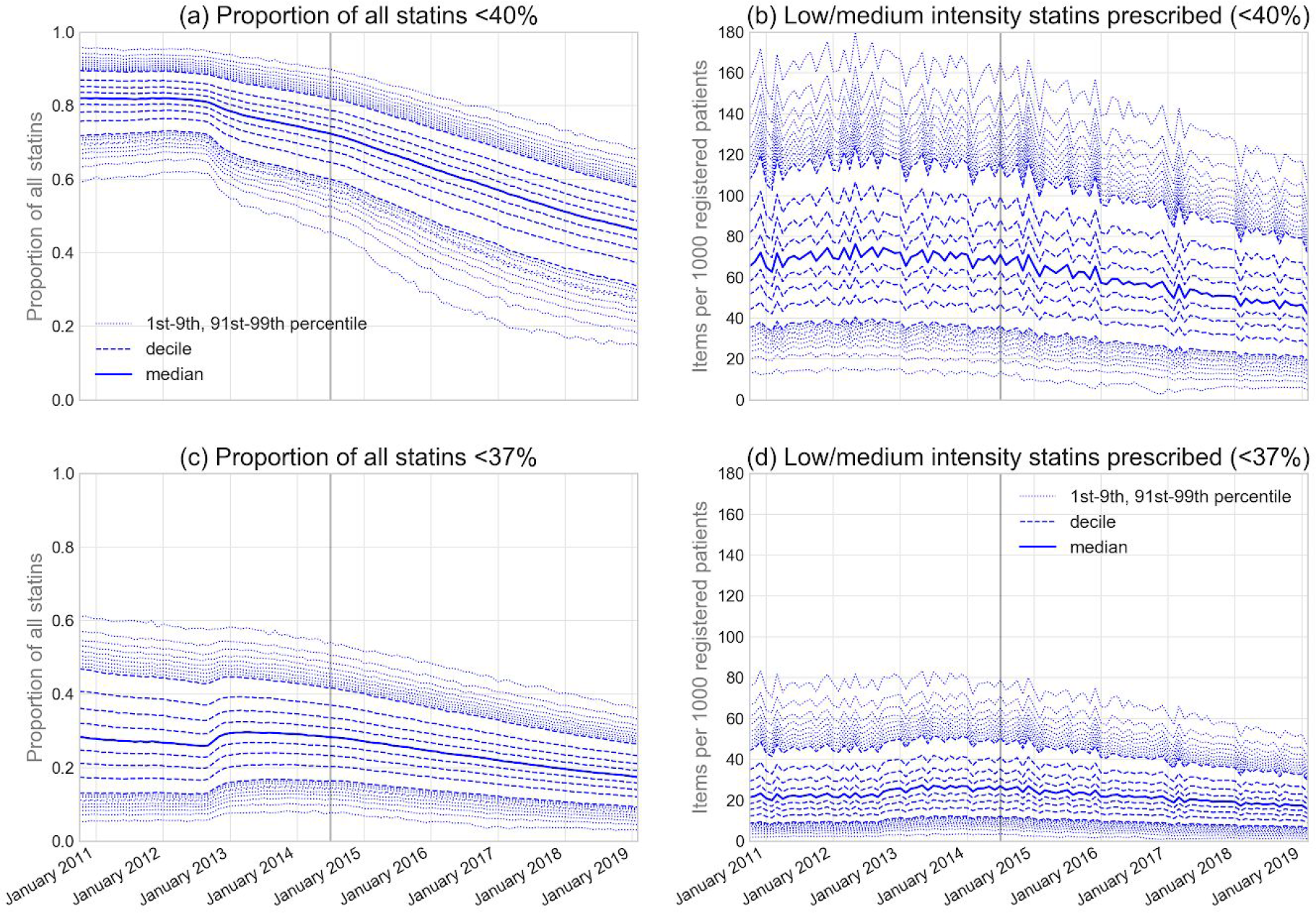
Monthly prescribing rates of low and medium-intensity statin across England’s practices from 2011-2019. (a, c) Proportion of all statin items under given intensity threshold and (b, d) Number of items under given intensity threshold prescribed per 1,000 registered patients. Solid line = median, dashed lines = deciles, light dotted lines = extreme percentiles (1-9th and 91-99th). Vertical line indicates release of NICE guidance, July 2014.

Nonetheless, in 2018, 10% of practices still prescribed more than 60% of statins in these low and medium-intensity forms, with only a small percentage of practices achieving 20% or less (Figure 3a). Compared to the proportion, the decline in absolute prescribing rate per 1000 population has been less pronounced (Figure 3b), and with very wide variety in performance: in 2018 10% of practices prescribed 25 or fewer per 1000 patients per month; while the top 10% prescribed at least 80. For comparison, the monthly prescribing rate for all statins interdecile range was 50 to 160 (eFigure 2a). For statins below the 37% threshold, the level of variation has narrowed, with interdecile range in the proportion prescribed reducing from almost 30% to less than 20% (Figure 3c).

### Practices that have changed quickly

Although the overall national change in the use of low and medium-intensity statins was slow, at 5.4 percentage point reduction in proportion per year (Figure 1b), our indicator saturation method for detecting practices with sudden rapid change found a large number of GP practices demonstrating very rapid changes in preferred treatment choice. For example, since 2014, there were 96 practices with a change of >5 percentage points per month amounting to a total change of >25 percentage points. These practices were distributed amongst 57 CCGs. Some examples of the practices with the quickest changes are illustrated in Figure 4.

**Figure 4:**
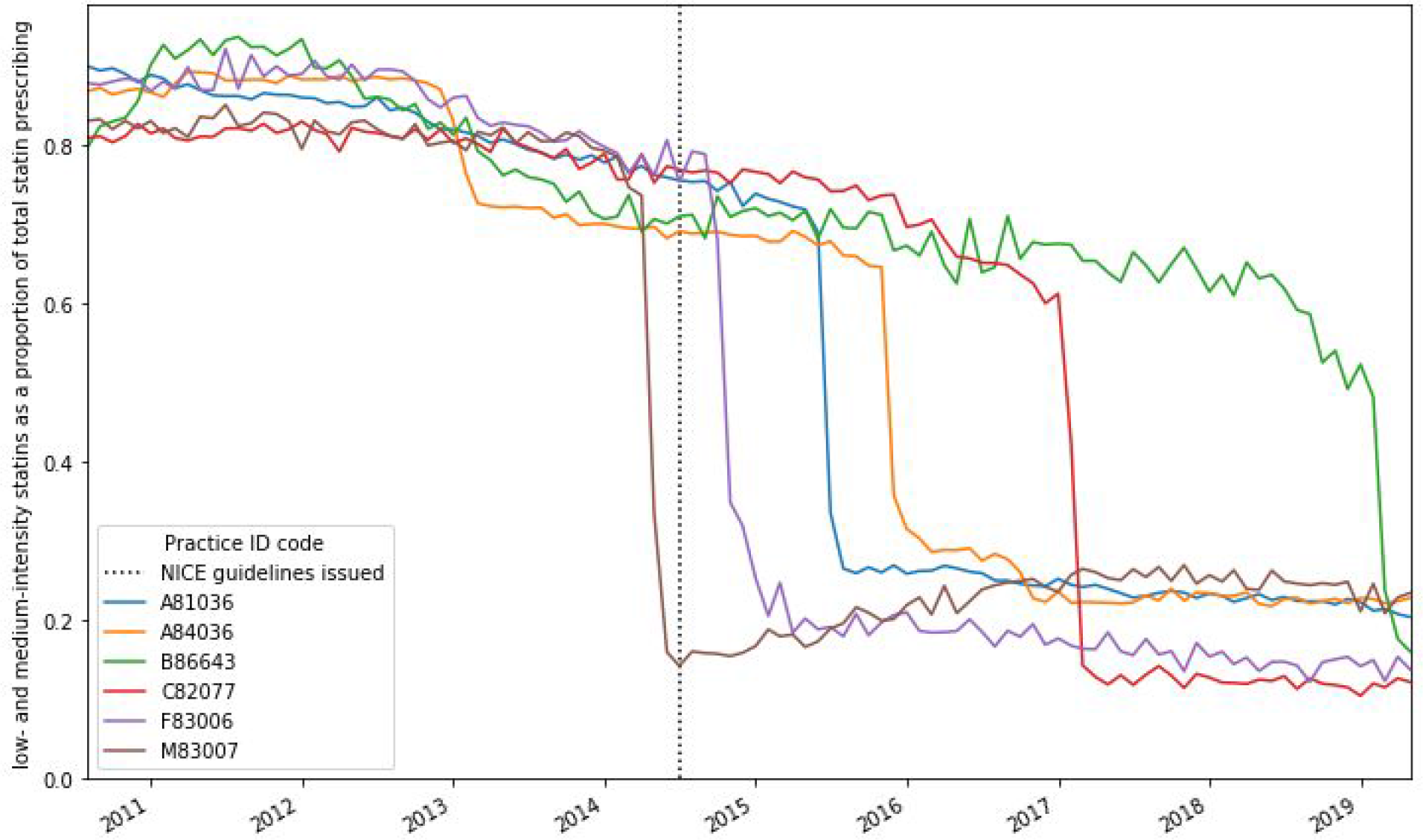
Examples of practices that have rapidly reduced their proportion of low- and medium-intensity statin (<40%) prescribing over the latest five years. Vertical line indicates release of NICE guidance, July 2014.

### Regression

We modelled the practice factors associated with the proportion of low- and medium-intensity statin prescribing in 2018 (Table 1). In the multivariable regression, very few of the variables had a substantial association with prescribing of low- and medium-intensity statins. The only meaningful association was with patient age, where practices with the highest proportion of patients over the age of 65 were slightly more likely to prescribe a greater proportion of low- and medium-intensity statins (multivariable odds ratio for youngest vs oldest: 1.22, 95% CI 1.17-1.28).

**Table 1.** Unadjusted and adjusted estimates for practice-level prescribing of low and medium-intensity statins as a proportion of all statins, from logistic regression analysis. CI = confidence interval, IMD = Index of Multiple Deprivation, QOF = Quality Outcomes Framework.

|  |  | Median<br>high<br>dose % | Univariable logistic<br>regression |  |  | Multivariable logistic<br>regression |  |  |
| --- | --- | --- | --- | --- | --- | --- | --- | --- |
|  |  |  | Odds<br>ratio | 95% CI |  | Odds<br>ratio | 95% CI |  |
| % of patients<br>over 65 | 0-10.8 | 0.16 | Ref |  |  | Ref |  |  |
|  | 10.8-15.4 | 0.18 | 1.22 | 1.18 | 1.26 | 1.13 | 1.09 | 1.16 |
|  | 15.4-18.9 | 0.19 | 1.28 | 1.24 | 1.32 | 1.16 | 1.12 | 1.20 |
|  | 18.9-22.7 | 0.20 | 1.31 | 1.27 | 1.35 | 1.18 | 1.14 | 1.23 |
|  | 22.7-89.8 | 0.20 | 1.31 | 1.27 | 1.36 | 1.22 | 1.17 | 1.28 |
| % with a long<br>term health<br>condition | 10.0-43.8 | 0.17 | Ref |  |  | Ref |  |  |
|  | 43.8-49.4 | 0.19 | 1.13 | 1.09 | 1.16 | 1.02 | 0.99 | 1.05 |
|  | 49.4-53.6 | 0.19 | 1.16 | 1.12 | 1.20 | 1.03 | 1.00 | 1.06 |
|  | 53.6-58.3 | 0.19 | 1.13 | 1.09 | 1.17 | 1.01 | 0.97 | 1.04 |
|  | 58.3-92.5 | 0.18 | 1.11 | 1.07 | 1.14 | 1.00 | 0.97 | 1.04 |
| Practice list<br>size (thousands) | 0-4.1 | 0.18 | Ref |  |  | Ref |  |  |
|  | 4.1-6.1 | 0.18 | 1.00 | 0.96 | 1.03 | 1.01 | 0.98 | 1.04 |
|  | 6.1-8.6 | 0.18 | 1.03 | 0.99 | 1.06 | 1.01 | 0.98 | 1.04 |
|  | 8.6-11.8 | 0.19 | 1.01 | 0.98 | 1.05 | 0.98 | 0.95 | 1.01 |
|  | 11.8-72.5 | 0.19 | 1.06 | 1.03 | 1.10 | 0.99 | 0.96 | 1.02 |
| Urban/rural | Urban major conurbation | 0.18 | Ref |  |  | Ref |  |  |
|  | Urban minor conurbation | 0.17 | 0.99 | 0.93 | 1.05 | 0.99 | 0.88 | 1.11 |
|  | Urban city and town | 0.20 | 1.15 | 1.13 | 1.18 | 1.02 | 0.97 | 1.07 |
|  | Rural town and fringe | 0.20 | 1.15 | 1.11 | 1.19 | 0.96 | 0.91 | 1.02 |
|  | Rural village and dispersed | 0.17 | 1.00 | 0.95 | 1.06 | 0.84 | 0.79 | 0.90 |
| IMD | Least deprived | 0.20 | Ref |  |  | Ref |  |  |
|  |  | 0.20 | 0.96 | 0.93 | 1.00 | 0.99 | 0.96 | 1.02 |
|  |  | 0.19 | 0.92 | 0.89 | 0.95 | 0.98 | 0.95 | 1.01 |
|  |  | 0.18 | 0.86 | 0.84 | 0.89 | 0.96 | 0.93 | 1.00 |
|  | Most deprived | 0.16 | 0.76 | 0.74 | 0.79 | 0.89 | 0.85 | 0.93 |
| QOF | 14-523 | 0.19 | Ref |  |  | Ref |  |  |
|  | 523-541 | 0.19 | 1.00 | 0.97 | 1.03 | 0.99 | 0.96 | 1.01 |
|  | 541-550 | 0.19 | 0.99 | 0.96 | 1.03 | 0.98 | 0.95 | 1.01 |
|  | 550-557 | 0.18 | 0.99 | 0.96 | 1.02 | 0.97 | 0.94 | 1.00 |
|  | 557-559 | 0.19 | 1.01 | 0.97 | 1.04 | 0.96 | 0.93 | 0.99 |

All other factors had odds ratios close to 1 (range 0.84 to 1.03). The CCG to which a practice belongs (as a random effect) was significantly associated with high-dose prescribing (p <0.0001) and accounted for 25.7% of the variation in low- and medium-intensity statin prescribing. 350 practices were excluded from the multivariable analysis due to missing data.

## Discussion

### Summary

English national guidance recommends the use of high-intensity statins. Breaches of this guidance in primary care are becoming less prevalent but remain extremely common, with substantial variation between practices. Using NICE criteria of treatments giving a 40% reduction in LDL cholesterol to identify breaches, low and medium-intensity statins fell from 80% of all statin prescriptions in 2011-12, to 45% in 2019, at a rate of 5.4 percentage points per year. The proportion of statins prescribed in formulations giving <37% LDL reduction (the more permissive criteria for breaching guidance) also fell, from a peak of 30% in 2013 to 18% in 2019. The NICE guidance released in 2014 had minimal impact on these trends. There was very substantial variation in prescribing behaviour between practices, with an interdecile range of 20-85% for NICE criteria, and 10-30% for the <37% LDL criteria. We found several examples of GP practices exhibiting a rapid change in prescribing behaviour towards greater guideline compliance, demonstrating that such changes are readily deliverable.

### Strengths and weaknesses

A major strength is that our data covers almost the entire population of England, thus minimising the potential for obtaining a biased sample. We used real prescribing and reimbursement data which are sourced from pharmacy claims and include all dispensed medication: these therefore did not need to rely on surrogate measures. Furthermore, these records are highly accurate, as this determines the flow of money from healthcare organisations to pharmacy businesses. While prescriptions issued by a hospital clinic or private practice or dispensed in hospital are not included in our data, the overwhelming majority of statin prescriptions are issued through a general practice. An additional strength of our study is that we examined both the NICE cut-off point of 40% reduction, and a more permissive cut-off of 37%, to account for the likely circumstance that GPs would prioritise changing the medication of patients whose current statin treatment substantially breaches current NICE guidance.

We used the proportion of statins in breach of guidance, rather than the absolute number of breaching prescriptions per head of local population, to account for confounding by indication, or variation in the number of people in each practice who are eligible for and treated with statins. There will be some situations where patients are prescribed lower intensity statins appropriately, as per the exceptions set out in the guidance: for example due to intolerance or perceived intolerance of higher doses [4]. However, such intolerance is relatively uncommon [17,18], and certainly less prevalent than the prescribing of low and medium-intensity statins identified in our data. Furthermore, it is very highly unlikely that the variation in prevalence of intolerance could be on the scale of the variation in prescription of low and medium-intensity statins we observed between practices, especially given the very high numbers involved. This could be assessed by interrogating richer electronic health record (EHR) data: however, in our clinical and analytic experience, concerns regarding statin intolerance are unlikely to be recorded consistently as structured data; and in any case there is no national EHR dataset covering all NHS GP practices in which to conduct such an analysis.

### Findings in Context

Our findings on the prevalence of high-intensity prescribing are consistent with previous work on smaller populations at single time-points in UK patient-level datasets. For example, it was found that prior to guideline release, 24% of CVD and high-risk patients, and 31% of patients with vascular disease, received high-intensity statins [6,7]. Among CVD patients initiating statins between 2010-2013, only around 2% were started on low-intensity statins (<29% LDL reduction), 74% “moderate” intensity (27-43% reduction, including atorvastatin 20mg) and 23% on high-intensity (>42% reduction) [19]. Consistent with this, our data showed approximately 70-75% of statins were low and medium-intensity in 2014. Our study also revealed that usage of low and medium-intensity statins was already dropping prior to release of the 2014 guidelines, and has continued to do so.

Our findings are mirrored by previous work from outside the English health system, although our approach covers a wider number of patients and clinical scenarios. In the US, similar guidelines were released in 2013 recommending a high-intensity (>50% LDL reduction) for many eligible patients, and moderate-intensity (30-50%) for some subgroups [20]. A study across 161 cardiology practices identified a subsequent small increase in higher-intensity statin use, continuing a pre-existing trend [21]. Further, another study including 140,000 US patients eligible for secondary prevention found that guideline adherence was strongly associated with geography, indicating that local policy or culture plays an important role [22]. Low levels of high-intensity statin usage have also been reported across Europe and worldwide, with a substantial proportion of patients not achieving target cholesterol levels [23,24].

Previous work has shown that doctors tend to respond rapidly to safety concerns around prescribing, while evidence-based guidelines have less impact, even when the prescribing advice is clear [25,26]. Our results support this, showing both a slow change in response to the 2014 guidelines on effective dosage, and a rapid change in late 2012 coinciding with an MHRA drug safety alert. This alert restricted the maximum dose of simvastatin to 20mg when used with two commonly prescribed medicines for CVD, amlodipine or diltiazem. Our data indicate that this caused a rapid reduction in simvastatin 40mg, with corresponding increases in simvastatin 20mg and atorvastatin. This followed an earlier alert on simvastatin 80mg due to potential side-effects as well as some contraindications for simvastatin [27].

Cost also appears to be a contributing factor in statin choice. Despite earlier evidence for the greater effectiveness of atorvastatin over simvastatin, it was not recommended widely in the NHS until its patent expired in 2012. Subsequently, the cost reductions from 2012 appear to have further increased atorvastatin use and reduced the use of simvastatin. The final high-intensity statin to come off patent was rosuvastatin in 2018. Its generally low usage over the study period is reflective of this high cost, and consistent with the NICE recommendations to use the statin with the lowest acquisition cost.

In a recent observational analysis, each 10% increase in intensity (eg, 30% to 40%) gave a hazard ratio of 0.90 (95% CI, 0.86-0.95) for cardiovascular events in CVD patients initiated on statins from 2010-2013 and followed up to the end of 2016 [19]. A combination of prescribing suboptimal statins and imperfect adherence (21% combined measure) led to 23.7 additional events per 1000 patient-years above the 48.3 predicted with perfect treatment (50% combined measure) [19].

### Policy Implications and Interpretation

The shortcomings in clinical practice that we have identified are likely to have a substantial negative impact on patient care. All patients with >10% ten-year risk of CVD are to be offered statins under NICE guidance [4]. If we conservatively assume an average 15% ten-year risk of a cardiovascular event for the total population taking statins then, as per the NICE risk calculator, with the recommended treatment of atorvastatin 20mg, their ten-year risk would reduce to 9% [28]. In other words, for every 1,000 patients treated with a high-intensity statin, 90 events would be expected over a ten-year period, compared to 150 if untreated, with 60 events prevented. Conservatively assuming a relative risk reduction of 33% for lower potency statins, only 50 events would be prevented in the same population of 1,000 patients at 15% ten year risk. We can therefore estimate that there will be ten avoidable cardiovascular events every ten years for every thousand patients inappropriately given a lower potency statin.

Our indicator-saturation change detection algorithm identified a large number of individual GP practices that exhibited a very rapid change in their statin prescribing to comply with national guidance on using higher potency statins, thus demonstrating that such changes can be - and are - readily implemented. Supporting this, we found the highest rates of optimal statin prescribing in central London and Bradford, which both have long-standing programmes to promote better statin prescribing, supported by bespoke local clinical guidance, software tools, and incentive schemes [29–31]. In Scotland, numerous interventions have been implemented to optimise statin prescribing, and it was reported that the proportion of atorvastatin prescribed in low doses decreased from 62.8% in 2001 to 19.7% in 2015 [32]. This all demonstrates that implementation of the NICE guidance to use higher intensity statins is readily achievable.

Having demonstrated that a substantial change in statin prescribing is necessary, important, and feasible, there is an outstanding question of how it can be achieved. We suggest that a national strategic approach is required, using data to identify outliers, and offering them feedback and targeted educational interventions. Audit and feedback alone has already been shown to be modestly effective at changing clinical practice [33]. We provide a free online data monitoring tool for anyone to see the current prescribing of high potency statins - and indeed all medicines - at any individual NHS general practice in England, through OpenPrescribing.net.

More broadly our findings and policy recommendations speak to a more general theme: the better use of existing low-cost techniques and interventions to improve clinical care. “Big data”, machine learning and artificial intelligence are commonly discussed as a future panacea, developing novel methods to identify and diagnose disease, identifying new treatments, personalising the use of older treatments, and delivering healthcare services in new ways; less commonly discussed is that we already have data and analytical techniques, readily available today, that are not being used to identify outliers, implement guidance and improve care. Similarly, statins are an effective intervention, at very low cost, and the most commonly prescribed class of medication in the UK; and yet they are often prescribed sub-optimally, as elsewhere in the world. If we cannot achieve concordance with the evidence among NHS clinicians for statins, the most commonly prescribed class of treatment in the UK, then there is a great deal of work to be done in using data and medicines to optimise patient outcomes.

### Future Research

We identify two areas for further research. Firstly, mixed methods research, using data such as ours to identify practices that breach guidance on optimal statin prescribing, and realist evaluation or other qualitative methods to understand the reasons for this breach. Secondly, the evaluation of interventions that aim to improve prescribing: whether low cost, such as delivering feedback on clinical practice; or high cost, such as targeted educational interventions for breaching clinicians.

### Summary

Wide variation in high intensity statin prescribing exists across England, contrary to national guidance. Some regions have effectively implemented action to achieve good compliance while others lag behind. A national strategic approach on using data and evidence to optimise care is needed.

## Data Availability

All data is shared openly online alongside all code for data management and analysis: https://github.com/ebmdatalab/statins-dose-paper.

## Acknowledgements

The authors appreciate the ongoing contributions of Seb Bacon, Peter Inglesby and Dave Evans to the OpenPrescribing databases and website which makes this work possible.

## Contributorship

HC extracted the data and carried out trends and variation analysis with input from BG and BM, and using code written by RC to classify statins by their intensity. AW carried out regression and indicator saturation with input from HC and BG. HC, AW, BM and BG designed the methodology. HC drafted and edited the manuscript with input from all other authors. BG is guarantor. The corresponding author attests that all listed authors meet authorship criteria and that no others meeting the criteria have been omitted.

